# Telomere length among Chinese oldest old

**DOI:** 10.1101/2022.07.26.22278044

**Authors:** Suey S.Y. Yeung, Xingyan Wang, Suk Ling Ma, Yangchao Chen, Stephen Kwok Wing Tsui, Nelson Leung Sang Tang, Jean Woo

## Abstract

**Introduction:** Telomere length (TL) is generally regarded as a biomarker of aging. TL, which is influenced by sociodemographic factors, has been shown to be inversely associated with morbidity. However, most studies were examined in the youngest old and whether the findings can be extended to the oldest old is less clear. This study examined TL and its associated factors in Chinese oldest old in Hong Kong.

**Methods:** Follow-up data collected after 14 years from the baseline of the Mr. and Ms. Osteoporosis cohort were analyzed. A structured interview on sociodemographic factors and physical measurement was conducted. Frailty and sarcopenia status were respectively determined by Fried’s criteria and Asian Working Group for Sarcopenia definition. TL was measured by a molecular inversion probe - quantitative PCR (MIP-qPCR) assay and expressed as telomere / a single copy reference gene (T/S) ratio. Adjusted binary logistic regressions were used to examine the associations between TL and the presence of multimorbidity, age-related diseases, frailty and sarcopenia.

**Results:** Among 555 participants (mean age 83.6±3.8 years, 41.3% women), the mean T/S ratio was 1.01±0.20. Males had lower T/S ratio (0.97±0.20) compared with females (1.07±0.18) (p<0.001). A lower education level was related to a longer TL (p=0.016). Being a current smoker was related to a shorter TL (p=0.007). TL was not significantly different across categories of age, subjective socioeconomic status, drinking status, physical activity level and body mass index (p>0.05). There were no associations between TL and the presence of multimorbidity, diabetes, stroke, cardiovascular diseases, cognitive impairment, frailty and sarcopenia.

**Conclusion:** Among Chinese oldest old, males had shorter TL compared with females. TL was not associated with age-related diseases, frailty and sarcopenia in this age group. TL may not be a biological marker of aging among the oldest old.

## Introduction

Over the last several decades, life expectancy has increased worldwide. Hong Kong has sustained the world’s highest life expectancy at birth since 2013. The latest data reported a life expectancy of 82.9 years for males and 88.0 years for females in Hong Kong [1]. Telomere length (TL), which has been extensively studied in relation to aging and longevity, may play a role in explaining extended longevity among many other factors.

Telomeres are repetitive DNA sequences at the end of chromosomes and serve as caps to preserve genomic integrity and stability by protecting linear DNA molecules from being recognized as double-stranded DNA breaks. TL is influenced by various genetic and non-genetic determinants such as demographic (age, sex), social and environmental factors (diet, smoking, alcohol use, body mass index, socioeconomic status and physical activity) [2-6]. However, most of the studies were conducted in the youngest old (below 75 years old). Studies related to TL beyond the age of 75 years were limited.

Telomeres shorten with each cell cycle, therefore, it has been suggested that telomere shortening is one of the hallmarks of aging [7]. Shorter TL was associated with poor survival and agerelated diseases including cardiovascular diseases, neurodegenerative diseases and diabetes in the youngest old [8,9]. However, these findings were inconsistent among the oldest old. The Leiden 85+ study showed that TL was not associated with age-related diseases [10]. Similarly, the Newcastle 85+ study showed that TL was not associated with multimorbidity, cognitive impairment and disability [11]. While TL was not associated with geriatric syndromes such as frailty and sarcopenia in the youngest old [12], whether this finding can be extended to the oldest old is less clear.

Currently, qPCR method was commonly used to assess the TL in epidemiology and population studies [13]. It is also widely known as the relative TL measurement in which TL is expressed as a T / S (telomere / a single copy reference gene) ratio. However, many problems have been described for this method. They include the stringent requirements of high quality DNA template, poor reproducibility, batch effect, etc [14-17]. Some research groups even suggested that previous reported association between TL and social factors might only represent a systemic bias instead of a real association [18]. A recent paper used the qPCR methods in the UK Biobank samples and found that variance due to technical factors was huge. Technical factors have a strong influence on the results include batch of primer, batch of enzyme, operator among others [19]. Some of the assay problems could be attributed to the use of a multiple-bases mismatch primer (for example the Tel-1b primer) in qPCR for amplification of the telomere repeat motif. Here, we designed a new relative TL measurement method which completely eliminated the use of all mismatch primers by using two molecular inversion probes (MIPs) and an additional ligation step. Instead of using the native telomere fragments of the sample DNA as the direct template of qPCR, ligation of exogenous MIPs targeting the telomere and another reference repeat motif are used as the template of qPCR. Therefore, this new relative method for TL which could be applied in epidemiology may be helpful to enrich the availability of methods for studying this important topic.

Molecular inversion probe (MIP) has been extensively used in molecular biology for almost 30 years and was first used as a localized DNA detection method and were called padlock probe. In that version of MIP, there was no gap between the 5’ and 3’ end of the MIP after hybridization to the targets [20]. Later, it was used as a method of genotyping single nucleotide polymorphisms (SNPs) with a single-base gap between the 5’ and 3’ end of the MIP [21]. In the recent next generation sequence era, MIP has been further developed to as capturing probe to enrich target templates for library preparation of high throughput sequencing. It is sometimes referred as connector inversion probe (CIP) if gap filling was used as a mean of target enrichment for sequencing. Here, we describe a novel application of MIP in measurement of length (or abundance) of a repeat motif, specifically we measured TL by quantification of ligated (circularized) MIP that had been hybridized to the target motif. And the longer the telomere, more MIP are accommodated and after ligation (circularization) they become templates for quantification. The relative TL is quantified in a reaction using 2 MIPs, one MIP for telomere motif and another MIP for a reference genomic motif. Efficiency corrected delta-delta CT values correlate with TL.

To the best of our knowledge, data on the TL measurements in the oldest old and especially in Asia is limited. It is important to fill this research gap given the rapid growth of the aging population. Using samples from the Osteoporotic Fractures in Men and Women (Mr. and Ms. Osteoporosis) study, we examined TL expressed as a T/S ratio and its associated factors among the Chinese oldest old in Hong Kong.

## Materials and Methods

### Study population

This study is based on the follow-up data collected 14 years after the baseline of the Mr. and Ms. Osteoporosis cohort. A detailed description of the study has been previously reported [22]. In brief, 4000 community-dwelling Chinese men and women aged ≥65 years participated in a health check between 2001 and 2003. Participants were recruited using a stratified sampling method to equally assign them into 65-69, 70-74 and 75+ age groups. They should have been able to walk or take public transport to the study site. Those who had had bilateral hip replacements or were not competent to give informed consent were excluded. Participants were invited to re-attend for a repeated health check between 2015 and 2017. For the present analysis, participants who discontinued the 14-year follow-up assessment (n=2939) and those who attended the assessment but without a valid TL measurement (n=506) were excluded. The final sample included in the analysis was 555. All participants gave their written informed consent to participate in the study. The study was conducted in compliance with the Declaration of Helsinki and was approved by the Clinical Research Ethics Committee of the Chinese University of Hong Kong.

### Characteristics of participants

A structured interview was conducted to collect information on sex, age, education level (primary or below vs. secondary or above), subjective socioeconomic status, smoking status (never/ex-smoker vs. current smoker), current drinker (no vs yes), physical activity level, body mass index (BMI) and medical history. Subjective socioeconomic status was assessed by asking participants to mark their self-perceived position on a drawing of an upright ladder with 10 rungs. The most undesirable state with respect to their standing in the community (community status ladder) and in Hong Kong (Hong Kong ladder) was indicated by the lowest rung and the most desirable state was indicated by the highest rung [23]. Physical activity level was assessed using the 12-item Physical Activity Scale for the Elderly (PASE) [24]. Participants reported the average number of hours per day spent in leisure, household, and occupational physical activities in the past week. Activity weights for each item were determined based on the amount of energy spent, and each item score was computed by multiplying the activity weight with daily activity frequency reported. A PASE score was computed by summing each item score. A higher PASE score represents a higher physical activity level. BMI was calculated as body weight (kg) divided by height (meters squared). Body weight was measured using the Physician Balance Bean Scale (Healthometer, Bridgeview, IL) with participants wearing light clothing. Height was measured using the Holtain Harpenden Standiometer (Holtain Ltd, Crosswell, UK). Low BMI was defined as <20.0 kg/m^2^.

A list of diseases was used to assess the number of chronic diseases, including diabetes, hyperthyroidism, hypothyroidism, stroke, Parkinson’s disease, heart attack, congestive heart failure, hypertension, chronic obstructive pulmonary disease and cancer. Participants were asked if a healthcare professional has even told them that they had or have each of the preceding diseases. Multimorbidity was defined as having two or more chronic diseases. Cognitive impairment was assessed by trained research staff using the Chinese version of the Mini-Mental State Examination (MMSE) [25]. The MMSE scores range from 0 to 30, with a higher score reflecting better cognitive function. Cognitive impairment was defined by a MMSE score of <24 points [26]. Frailty status was defined using the 5-item Cardiovascular Health Study (CHS) frailty phenotype: self-rated exhaustion, weakness (grip strength), slow walking speed, low physical activity and unintentional weight loss [27].

The corresponding variables used in this study for the construction of the CHS score were a little of time over the past 4 weeks feel a lot of energy, handgrip strength ≤first quintile, 6-meter walking speed ≤first quintile, PASE score ≤first quintile, and weight loss ≥5% in the past 6 months. The total score ranges from 0 to 5. A score of 1 to 2 represents a pre-frail state, and a score of ≥3 represents frailty. Sarcopenia was defined using the Asia Working Group for Sarcopenia (AWGS) [28]. Participants who had low muscle mass and low muscle strength or low physical performance were defined as having sarcopenia. Muscle mass was measured using the dual-energy X-ray absorptiometry and total appendicular skeletal muscle mass (ASM) was calculated. Low muscle mass was defined as ASM/height^2^ <7.0 kg/m^2^ for males and <5.4 kg/m^2^ for females. Low muscle strength was defined as handgrip strength of <26 kg in males and <18 kg in females. Low physical performance was defined as a gait speed of <0.8 m/s using the 6-meter walking test at the usual pace [28].

### Telomere length measurements by MIP-qPCR

Two MIPs are used, one binds to telomere repeat motif (GGGTTA) and another binds to a common genomic repeat motif such as 4 bp or 5 bp repeats. The 4-bp ATGG repeat motif was used as the target of genomic repeat in this study. The details of DNA sequences and protocol are described in Supplementary Material. The length of the telomere determines the extent of availability of binding sites for the telomere specific MIPs, therefore also related to the abundance of circularized telomere MIP after ligase reaction (as shown in Fig. 1). On the other hand, the availability of genomic repeat motif represents the amount of template DNA added into the ligase reaction as amount of genomic repeat motif (e.g. ATGG) is fairly stable per copy of eukaryotic genome [29]. After a ligase reaction, the two types of circularized MIP are quantified. Here, we used quantitative real-time PCR. However, other quantification methods like digital PCR could be used as well. The ratio of the amount of circularized telomere MIP to that of genomic repeat MIP is proportional to telomere length. Like the original relative TL qPCR method, telomere length expressed as T/S ratio were obtained by efficiency corrected delta-delta method. In addition, calibration with samples of given telomere length could also be carried out to convert the T/S ratio to telomere length in kbp unit. Two calibrators with given TL (6.9 and 8.1 kbp) measured by Southern blot were used as calibrators which enable conversion of the ratio to telomere length in term of the unit of kbp.

**Fig. 1.**
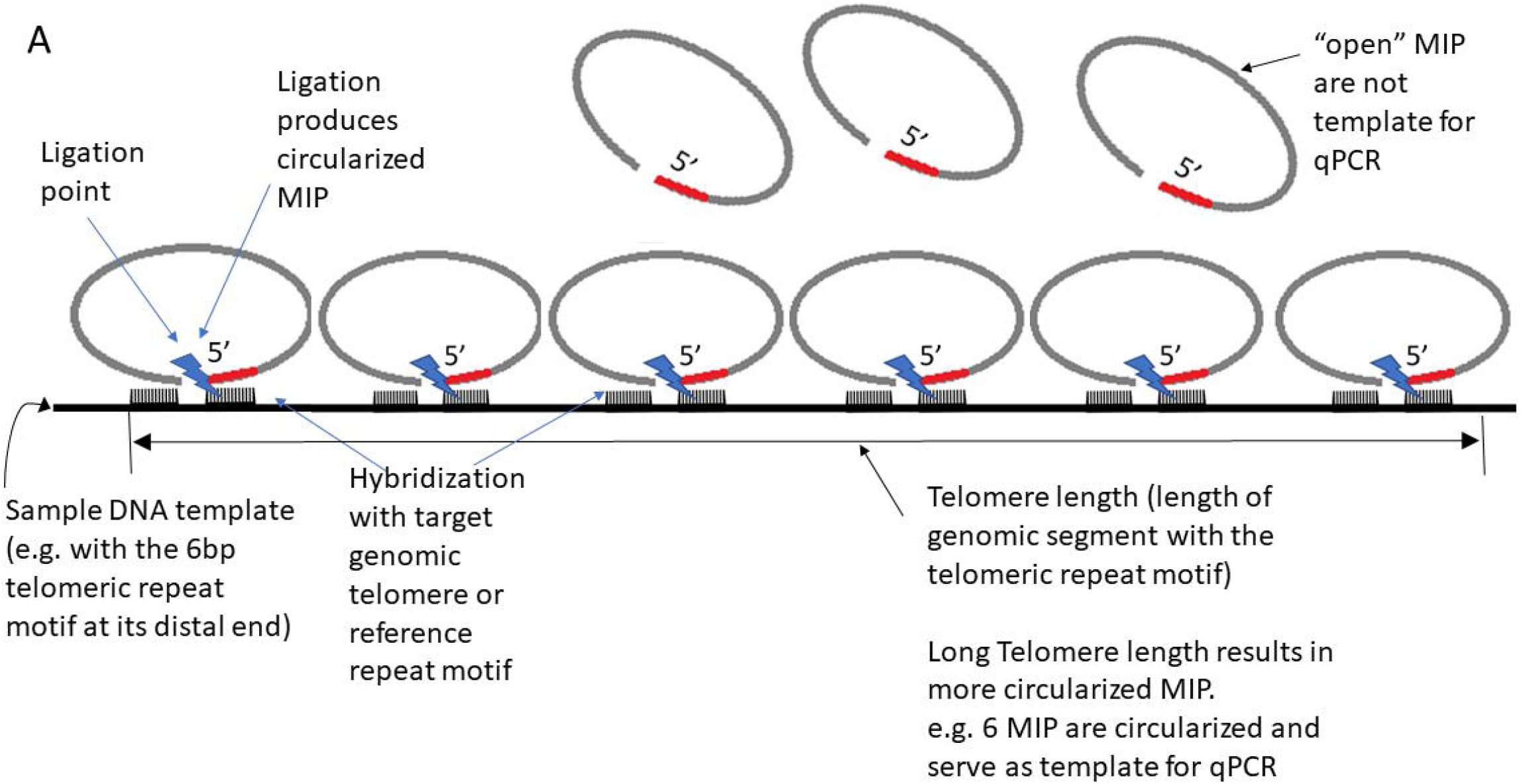
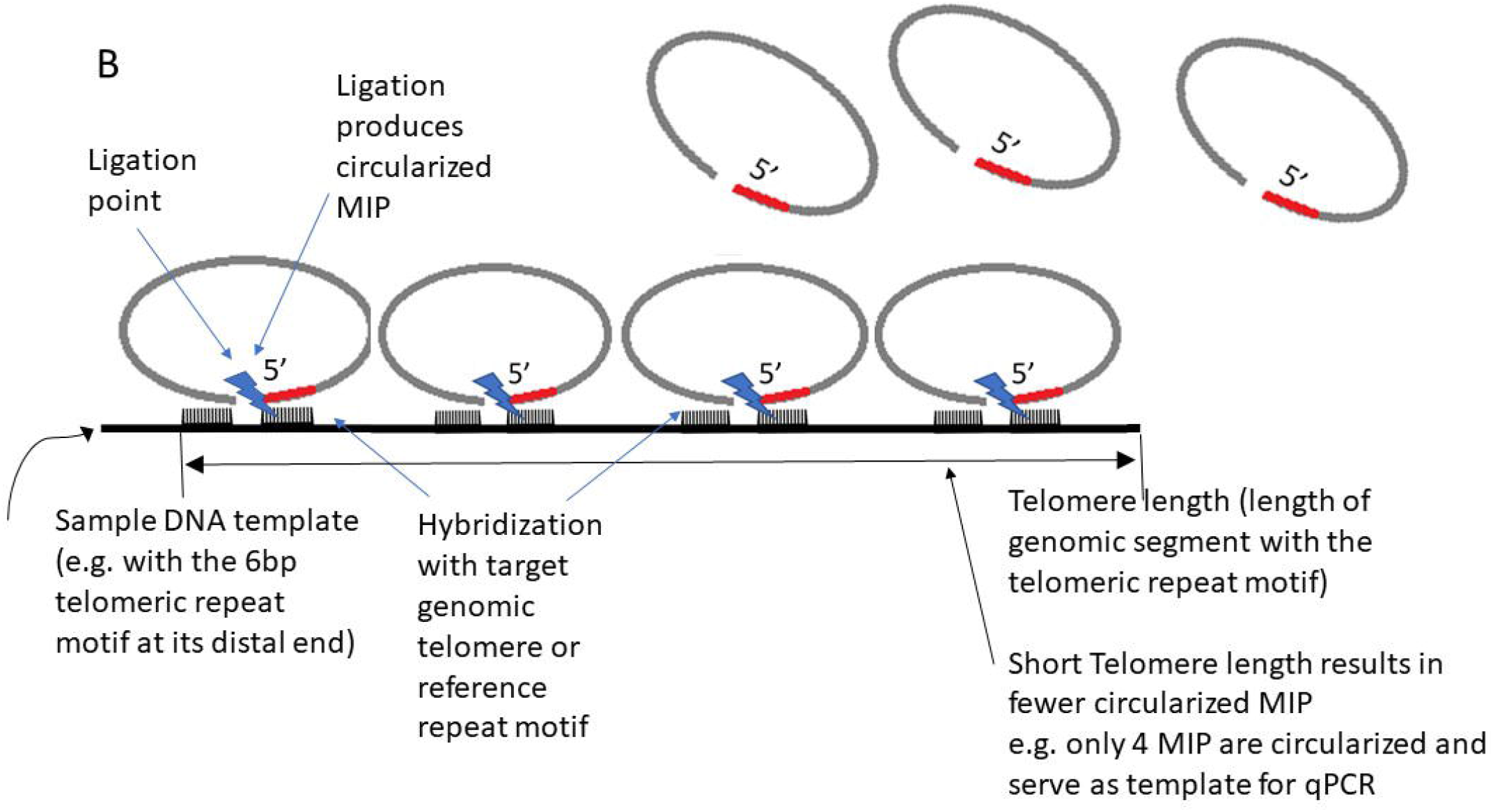
(A) It represents a sample with a long telomere and (B) represents another sample with a short telomere. Template DNA of the sample showing the region with telomere repeats are shown as the horizontal line. The length of the horizontal line represents telomere length which is longer in (A). Open molecular inversion probes (MIPs) are added to the samples. Both 5’ and 3’ ends of MIP hybridise to the target motif. Then hybridised MIPs are circularised by ligation reaction. These circularised MIPs are then used as template for qPCR to quantify the abundance of circularised MIP. Longer telomere (A) circularises more MIP.

DNA was extracted from blood samples using commercial DNA extraction kit according to the manufacturer’s instruction. The MIP for telomere consists of the telomere repeat sequence (GGGTTA) at both 5’ and 3’ ends connected by linker sequences. Another MIP is used to target a common 4-bp tandem repeat sequence (ATGG) for normalization as roughly the same abundance of such 4-bp motifs are present in each eukaryotic cell. Both MIPs for telomere and 4-bp tandem repeat sequence were 5’-phosphorylated and they were incubated overnight at 50°C with the DNA for hybridization. Gap filling and ligation was performed by the addition of Ampligase (EpiCentre), T4 polymerase (Roche) in the presence of 10mM of only dATP and dGTP, and incubated at 37°C for 30 minutes and inactivated at 95°C for 10 minutes. This ligase reaction circularizes those MIPs that have been hybridized to their respective target motifs.

To quantify the total number of circularized MIPs of the 2 types of repeat motifs, duplex quantitative PCR using specific primers to each type of MIP were performed. These primer pairs were designed specific to PCR amplify circularized telomere and 4-bp tandem repeat MIPs respectively and PCR does not proceed with non-circularized (open) MIPs. A 5’ GC rich tail was added to all primers. In addition, two Taqman probes containing the respective repeat sequence (GGGTTA or ATGG) were used in qPCR to differentiate the circularized template of the 2 types of MIP probes in a duplex qPCR performed in the same tube. Delta-delta CT with correction of PCR efficiency were calculated. That is delta Ct of each sample subtracted that of the reference sample to calculate the delta-delta Ct value. Based on the observed qPCR efficiencies of the 2 reactions, the ratio of telomere motif ligation points to genomic 4bp motif ligation points could be calculated as the efficiency corrected delta-delta Ct method. This efficiency corrected delta-delta Ct ratio of the 2 motifs of each sample is used as a biomarker of telomere length (T/S ratio). And average values was obtained from duplicated reactions for each sample.

In addition, results of TL in kbp unit can be obtained by calibration of T/S ratio against known TL of the calibrator samples. Efficiency corrected delta-delta Ct ratios of individual study samples were regressed onto the calibration line to provide the telomere length in the unit of kbp.

### Statistical analysis

Data are presented as numbers (percentages) and mean±standard deviation (SD). The differences in T/S ratio across categories of sociodemographic factors were examined using analysis of variance (ANOVA) or independent t-test, where appropriate. The associations between TL and each health outcome (the presence of multimorbidity, diabetes, stroke, cardiovascular diseases, cognitive impairment, frailty and sarcopenia) were examined using binary logistic regression models adjusting for covariates. Data are presented as odds ratio (OR) and 95% confidence intervals (CI).

All the analyses were stratified by sex. The analyses were performed using the Statistical Package for the Social Science (version 26.0, Chicago, IL, USA). Results were considered statistically significant when *p*<0.05.

## Results

### Characteristics of participants

Table 1 shows the descriptive characteristics of the 555 participants. The mean age was 83.6±3.8 years, and 41.3% were females. Most of the participants completed primary school or below (63.4%), were non-current smokers (97.7%) and had normal cognitive function (75.7%). The mean T/S ratio of Chinese oldest old was 1.01±0.2.

In Chinese oldest old, males had significantly shorter TL (T/S ratio 0.97±0.20) compared with females (T/S ratio 1.07±0.18) (p<0.001). Those who completed primary school or below (T/S ratio 1.03±0.19) had higher TL compared with those who completed secondary education or above (T/S ratio 0.98±0.21) (p=0.016). Being a non-current smoker (T/S ratio 1.01±0.20) had significantly higher TL compared with current smoker (T/S ratio 0.86±0.14) (p=0.004). TL was not significantly different across categories of age, subjective socioeconomic status, drinking status, physical activity level and BMI (p>0.05). There were no significant differences in TL between those with or without multimorbidity, diabetes, stroke, cardiovascular diseases, cognitive impairment, frailty and sarcopenia (p>0.05). Figure 2 shows the graphic representation of T/S ratio by age group for males and females.

**Fig. 2.**
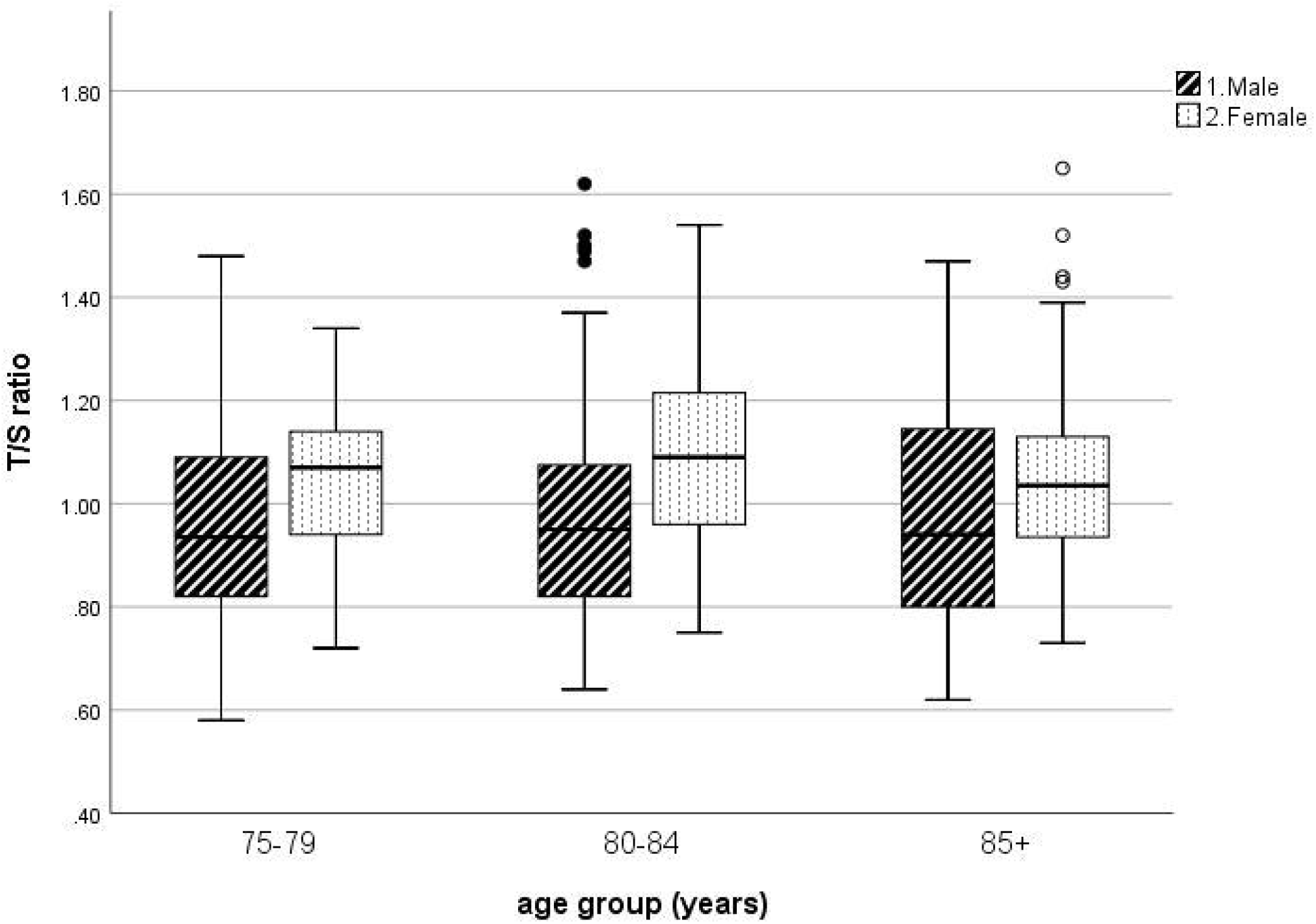
Telomere length (T/S ratio) by age group for males and females.

Table 2 shows the associations between TL and prevalent frailty and sarcopenia status. In crude and fully adjusted models, TL was not associated with prevalent frailty (adjusted OR for males 0.55, 95% CI 0.10-3.06, p=0.490; adjusted OR for females 0.19, 95% CI 0.02-1.51, p=0.117) and sarcopenia (adjusted OR for males 4.07, 95% CI 0.96-17.3, p=0.057; adjusted OR for females 0.74, 95% CI 0.10-5.62, p=0.769). No associations between TL and the presence of multimorbidity, diabetes, stroke, cardiovascular diseases and cognitive impairment were found in crude and adjusted models (p>0.05, data not shown).

## Discussion

The present study found a mean T/S ratio of 1.01 among Chinese oldest old (mean age of 83.6 years), with higher TL observed in males compared with females. TL was longer for those with lower education levels and among non-current smokers. There were no differences in TL across categories of other factors being investigated including age, subjective socioeconomic status, smoking status, physical activity level and BMI. TL was not associated with multimorbidity, diabetes, stroke, cardiovascular diseases, cognitive impairment, frailty and sarcopenia in Chinese oldest old.

To the best of our knowledge, this was the first study to investigate TL and its associated factors among the Chinese oldest old. The mean T/S ratio in our participants was higher compared with older adults in China (mean age 61 years, T/S ratio 0.67), US (mean age 71 years, T/S ratio 0.90) and Finland (mean age 71 years, T/S ratio 0.86) [30-32], but lower than Australian older adults (mean age 77 years, T/S ratio 1.18) [33]. These discrepancies might be explained by the different mean age, methods to measure TL or other sociodemographic factors. Consistent with the literature and the baseline analysis of the same cohort, we found that males had a shorter TL compared with females [5,34]. The sex-difference in TL could potentially be explained by the effect of estrogens. Estrogen level is associated with telomerase activity via telomerase reverse transcriptase (hTERT) gene expression, while androgen appears to be a negative regulator of telomerase in normal prostate tissue [35]. We found that oldest old with lower education levels had longer TL. A possible explanation is that those with lower educational levels worked for lower-level and physically demanding occupations, such that they were more physically healthy and experienced less work stress compared with those with higher education levels and worked for higher-level occupations. In line with the findings of a meta-analysis on smoking status and TL, we found that current smoker had a shorter TL than non- or former smokers [2]. Harmful chemicals in cigarettes may induce oxidative stress and inflammation, resulting in damage to the telomeric DNA and shortened telomeres. Most of the sociodemographic factors of TL in the younger age group do not appear in the oldest old. This suggests that other factors like genetics may have more influence on TL in the oldest old. Data of 287 twin pairs aged 73 to 95 years from the Danish Twin Registry showed that TL was moderately heritable, suggesting genetic control of individual TL [36].

In contrast with some studies which found that shorter TL was associated with diabetes, stroke, cardiovascular diseases, cognitive impairment, frailty and sarcopenia in the younger age group (up to a mean age of 75 years) [9,37-39], our study did not find such associations in the oldest old. Our findings were in line with the Leiden 85-plus study which showed that TL was not associated with a history of cardiovascular diseases, hypertension, diabetes, cognitive impairment and dementia in the oldest old. TL in this advanced age group was found to either shorten or lengthen by a considerate amount within a few years. The authors suggested that TL is not a predictive indicator for age-related morbidity, especially among the oldest old, due to a high degree of TL instability in this age group [10]. Similarly, the Newcastle 85+ study showed that TL was not associated with the presence of multimorbidity, cognitive impairment and disability [11]. In a large cohort of the oldest old in Switzerland (mean age 85 years), no significant difference in TL was found between cognitively normal individuals and those with dementia or mild cognitive impairment [40]. No study has examined the association between TL and frailty and sarcopenia in the oldest old. However, the lack of such association in the present study is compatible with a systematic review and meta-analysis in the younger age group [12]. Findings from our study appear to support that TL should not be used unconditionally as a biomarker of the human aging process in older populations [10]. Prospective studies are warranted to provide further information on TL attrition and its role in aging in the oldest old.

The strengths of this study include the use of data from a relatively large sample of Chinese oldest old and a series of socioeconomic and demographic factors and health outcomes. Several limitations of the present study should be noted. The cross-sectional design restricts us from drawing conclusions about causal relationships. Those with diseases may have passed away earlier, therefore, fewer associations with diseases may be observed. The reliance on self-report of health conditions may be another limitation in the current study. Furthermore, factors accumulated over the life course, which may influence TL, were not captured. Dietary data at 14-year follow-up was not available. However, we found that baseline dietary intake in terms of food groups and dietary patterns were not associated with TL at 14-year follow-up (data not shown). In addition, our results may not be generalizable to other populations as our participants were volunteers who were more likely to be health-conscious and have better health status than the general population.

To conclude, among Chinese oldest old, males had lower TL compared with females. A lower education level and being a non-current smoker was related to higher TL. TL was not associated with the presence of multimorbidity, age-related diseases, frailty and sarcopenia in this age group. It appears that TL should not be considered a biological marker of aging among the oldest old.

## Data Availability

The datasets generated and/or analyzed during the current study are available from the corresponding author on reasonable request.

## Statements of Ethics

The subjects have given their written informed consent and that the study protocol was approved by the Clinical Research Ethics Committee of the Chinese University of Hong Kong (CRE 2003.102).

## Conflict of Interest Statement

The method of telomere length measurement is pending patent proceedings.

## Funding Sources

This work was supported by the Hong Kong Jockey Club Charities Trust. We thank the generous donation of Ms. Therese Pei Fong Chow. The funders had no role in the design, methods, subject recruitment, data collections, analysis or preparation of this article.

## Author Contributions

Authors contributed to this work are as follows: S.S.Y.Y. performed the statistical analysis, interpreted the results, and wrote the manuscript; X.W. performed the experiments; S.L.M. prepared the manuscript; Y.C. and S.K.W.T. performed statistical analysis; N.L.S.T. contributed to discussions about the results and critically revised the manuscript; J.W. designed the research, contributed to discussions about the results and critically revised the manuscript. All the authors read and approved the final version submitted.

## Notes

### Author Declarations

Clinical Research Ethics Committee of the Chinese University of Hong Kong gave ethical approval for this work

## References

1 Centre for Health Protection: Life Expectancy at Birth (Male and Female), 1971 - 2020. Hong Kong, Department of Health, 2021 [cited 2022 Apr 29]. Available from: https://www.chp.gov.hk/en/statistics/data/10/27/111.html.

2 Astuti Y, Wardhana A, Watkins J, Wulaningsih W, Network PR. Cigarette smoking and telomere length: A systematic review of 84 studies and meta-analysis. Environ Res. 2017;158:480–489.

3 Lin X, Zhou J, Dong B. Effect of different levels of exercise on telomere length: A systematic review and meta-analysis. J Rehabil Med. 2019;51:473–478.

4 Robertson T, Batty GD, Der G, Fenton C, Shiels PG, Benzeval M. Is socioeconomic status associated with biological aging as measured by telomere length? Epidemiol Rev. 2013;35:98–111.

5 Gardner M, Bann D, Wiley L, Cooper R, Hardy R, Nitsch D, et al. Gender and telomere length: systematic review and meta-analysis. Exp Gerontol. 2014;51:15–27.

6 Muezzinler A, Zaineddin AK, Brenner H. Body mass index and leukocyte telomere length in adults: a systematic review and meta-analysis. Obes Rev. 2014;15:192–201.

7 Lopez-Otin C, Blasco MA, Partridge L, Serrano M, Kroemer G. The hallmarks of aging. Cell. 2013;153:1194–1217.

8 Wang Q, Zhan Y, Pedersen NL, Fang F, Hagg S. Telomere length and all-cause mortality: A meta-analysis. Ageing Res Rev. 2018;48:11–20.

9 Smith L, Luchini C, Demurtas J, Soysal P, Stubbs B, Hamer M, et al. Telomere length and health outcomes: An umbrella review of systematic reviews and meta-analyses of observational studies. Ageing Res Rev. 2019;51:1–10.

10 Martin-Ruiz CM, Gussekloo J, van Heemst D, von Zglinicki T, Westendorp RG. Telomere length in white blood cells is not associated with morbidity or mortality in the oldest old: a population-based study. Aging Cell. 2005;4:287–290.

11 Martin-Ruiz C, Jagger C, Kingston A, Collerton J, Catt M, Davies K, et al. Assessment of a large panel of candidate biomarkers of ageing in the Newcastle 85+ study. Mech Ageing Dev. 2011;132:496–502.

12 Lorenzi M, Bonassi S, Lorenzi T, Giovannini S, Bernabei R, Onder G. A review of telomere length in sarcopenia and frailty. Biogerontology. 2018;19:209–221.

13 Cawthon RM. Telomere measurement by quantitative PCR. Nucleic Acids Res. 2002;30:e47.

14 Dagnall CL, Hicks B, Teshome K, Hutchinson AA, Gadalla SM, Khincha PP, et al. Effect of pre-analytic variables on the reproducibility of qPCR relative telomere length measurement. PLOS ONE. 2017;12:e0184098.

15 Mensà E, Latini S, Ramini D, Storci G, Bonafè M, Olivieri F. The telomere world and aging: Analytical challenges and future perspectives. Ageing Research Reviews. 2019;50:27–42.

16 Eisenberg DTA, Kuzawa CW, Hayes MG. Improving qPCR telomere length assays: Controlling for well position effects increases statistical power. Am J Hum Biol. 2015;27:570–575.

17 Goldman EA, Eick GN, Compton D, Kowal P, Snodgrass JJ, Eisenberg DTA, et al. Evaluating minimally invasive sample collection methods for telomere length measurement. Am J Hum Biol. 2018;30

18 Geronimus AT, Bound J, Mitchell C, Martinez-Cardoso A, Evans L, Hughes L, et al. Coming up short: Comparing venous blood, dried blood spots & saliva samples for measuring telomere length in health equity research. PLOS ONE. 2021;16:e0255237.

19 Codd V, Denniff M, Swinfield C, Warner SC, Papakonstantinou M, Sheth S, et al. Measurement and initial characterization of leukocyte telomere length in 474,074 participants in UK Biobank. Nat Aging. 2022;2:170–179.

20 Nilsson M, Malmgren H, Samiotaki M, Kwiatkowski M, Chowdhary BP, Landegren U. Padlock probes: circularizing oligonucleotides for localized DNA detection. Science (New York, NY). 1994;265:2085–2088.

21 Hardenbol P, Banér J, Jain M, Nilsson M, Namsaraev EA, Karlin-Neumann GA, et al. Multiplexed genotyping with sequence-tagged molecular inversion probes. Nat Biotechnol. 2003;21:673–678.

22 Wong SY, Kwok T, Woo J, Lynn H, Griffith JF, Leung J, et al. Bone mineral density and the risk of peripheral arterial disease in men and women: results from Mr. and Ms Os, Hong Kong. Osteoporos Int. 2005;16:1933–1938.

23 Adler NE, Epel ES, Castellazzo G, Ickovics JR. Relationship of subjective and objective social status with psychological and physiological functioning: preliminary data in healthy white women. Health Psychol. 2000;19:586–592.

24 Washburn RA, Smith KW, Jette AM, Janney CA. The Physical Activity Scale for the Elderly (PASE): development and evaluation. J Clin Epidemiol. 1993;46:153–162.

25 Folstein MF, Folstein SE, McHugh PR. “Mini-mental state”. A practical method for grading the cognitive state of patients for the clinician. J Psychiatr Res. 1975;12:189–198.

26 Creavin ST, Wisniewski S, Noel-Storr AH, Trevelyan CM, Hampton T, Rayment D, et al. Mini-Mental State Examination (MMSE) for the detection of dementia in clinically unevaluated people aged 65 and over in community and primary care populations. Cochrane Database Syst Rev. 2016:CD011145.

27 Fried LP, Tangen CM, Walston J, Newman AB, Hirsch C, Gottdiener J, et al. Frailty in older adults: evidence for a phenotype. J Gerontol A Biol Sci Med Sci. 2001;56:M146–156.

28 Chen LK, Liu LK, Woo J, Assantachai P, Auyeung TW, Bahyah KS, et al. Sarcopenia in Asia: consensus report of the Asian Working Group for Sarcopenia. J Am Med Dir Assoc. 2014;15:95–101.

29 Gymrek M, Willems T, Guilmatre A, Zeng H, Markus B, Georgiev S, et al. Abundant contribution of short tandem repeats to gene expression variation in humans. Nat Genet. 2016;48:22–29.

30 Haapanen MJ, Perala MM, Salonen MK, Guzzardi MA, Iozzo P, Kajantie E, et al. Telomere length and frailty: The Helsinki Birth Cohort Study. J Am Med Dir Assoc. 2018;19:658–662.

31 Huang Z, Liu C, Ruan Y, Guo Y, Sun S, Shi Y, et al. Dynamics of leukocyte telomere length in adults aged 50 and older: a longitudinal population-based cohort study. Geroscience. 2021;43:645–654.

32 Bello GA, Chiu Y-HM, Dumancas GG. Association of a biomarker-based frailty index with telomere length in older US adults: Findings from NHANES 1999-2002. bioRxiv. 2017:191023.

33 Yeap BB, Hui J, Knuiman MW, Flicker L, Divitini ML, Arscott GM, et al. U-shaped relationship of leukocyte telomere length with all-cause and cancer-related mortality in older men. J Gerontol A Biol Sci Med Sci. 2021;76:164–171.

34 Woo J, Tang NL, Suen E, Leung JC, Leung PC. Telomeres and frailty. Mech Ageing Dev. 2008;129:642–648.

35 Bayne S, Liu JP. Hormones and growth factors regulate telomerase activity in ageing and cancer. Mol Cell Endocrinol. 2005;240:11–22.

36 Bischoff C, Graakjaer J, Petersen HC, Hjelmborg J, Vaupel JW, Bohr V, et al. The heritability of telomere length among the elderly and oldest-old. Twin Res Hum Genet. 2005;8:433–439.

37 Martinez-Ezquerro JD, Rodriguez-Castaneda A, Ortiz-Ramirez M, Sanchez-Garcia S, Rosas-Vargas H, Sanchez-Arenas R, et al. Oxidative stress, telomere length, and frailty in an old age population. Rev Invest Clin. 2019;71:393–401.

38 Haycock PC, Heydon EE, Kaptoge S, Butterworth AS, Thompson A, Willeit P. Leucocyte telomere length and risk of cardiovascular disease: systematic review and meta-analysis. BMJ. 2014;349:g4227.

39 Marzetti E, Lorenzi M, Antocicco M, Bonassi S, Celi M, Mastropaolo S, et al. Shorter telomeres in peripheral blood mononuclear cells from older persons with sarcopenia: results from an exploratory study. Front Aging Neurosci. 2014;6:233.

40 Zekry D, Herrmann FR, Irminger-Finger I, Ortolan L, Genet C, Vitale AM, et al. Telomere length is not predictive of dementia or MCI conversion in the oldest old. Neurobiol Aging. 2010;31:719–720.

