## Supplementary Material TL 14y Os for "Telomere length among Chinese oldest old"

**Telomere MIP protocol for samples-2021**

1. MIP step 1 （hybridization step）

MIP sequence: (Yellow color sequence binds to the target motif, underlined sequence are binding location of primers in qPCR)

1) **Tel:** MIP_ShortTelo2021:

/5Phos/GGTTAGGGTTAGGGTTACAGGCCAAGTAAAACGCACTTAGTGAGACCCCTTTAGCGTAAGAACAATGGTTAGGGTTAGGGTTA

2) **4bp motif (ATGG)**: ShortMIP4bp:

/5Phos/GATGGATGGATGGATGGCTTCAGCTTCCCGATCCGACGGTTAGGTTCACACAGGAAACAGCTATGACTGGATGGATGGATGGAT

3) Protocol (15:15) :

| DNA (10ng/ul) | 8ul |
| --- | --- |
| Amp ligase buffer (10x) | 2.5ul |
| MIP_ShortTelo2021 (10nM) | 3ul |
| ShortMIP4bp2019 (10nM) | 3ul |
| H_2_O | 8.5ul |
| Total volume | 25ul |

Hybridization protocol

95℃	10min

72℃	1min, ramp rate 0.1℃/s

50℃	5min, ramp rate 0.1℃/s

Goto 2, 10X

50℃	16 hours, ramp rate 0.1℃/s

2. MIP step 2 (gap filling and ligation step to circularise MIP that have been hybridized to their target motifs)

Put the MIP product (pdt) from MIP step 1 to room temperature.

Mix 5ul MIP pdt of MIP step 1 with the following mixture:

| Amp ligase (5U/ul) | 0.8ul |
| --- | --- |
| T4 polymerase (3U/ul) | 0.15ul |
| dGTP (10mM) | 0.5ul |
| dATP(10mM) | 0.5ul |
| BSA (100X) | 0.2ul |
| Amp ligase buffer (10X) | 1.5ul |
| H_2_O | 11.35ul |
| Total volume | 15ul |

Temperature protocol for gap filling and ligation

| 37℃ | 30min |
| --- | --- |
| 45℃ | 5min |
| 95℃ | 10min |
| 4℃ | keep |

3. qPCR (quantification of circularized products)

Primer sets: (all with a 5’ GC tail, underlined sequence are binding location of primers against MIPs in qPCR)

| Short_telo2021F | gccgctgac CC CCT TTA GCG TAA GAA CAA TGG |
| --- | --- |
| Short_telo2021R | gacctcgtgct GTG CGT TTT ACT TGG CCT |
| longGC_M13_F | ggcgcatggc TCA CAC AGG AAA CAG CTA TGA C |
| longGC_Link-R_ATGG | gcatggcgca ATC GGG AAG CTG AAG CCA TCCAT |

Taqman probes

| Taqman for Telo | /5' 6-FAM/TA GGG TTA GGG TTA GGG TTA GGGT/3' lowa Black FQ/ |
| --- | --- |
| Taqman for ATGG | /5' 6-HEX/ATCCATCCATCCATCCATCCgTC/Blackhole/ |

Protocol:

| Roche probe master mix (2X) | 7.5ul |
| --- | --- |
| Short_telo2021F (20uM) | 0.375ul |
| Short_telo2021R (20uM) | 0.375ul |
| longGC_M13_F (20uM) | 0.375ul |
| longGC_Link-R_ATGG (20uM) | 0.375ul |
| Taqman for Telo (10uM) | 0.3ul |
| Taqman for ATGG (10uM) | 0.3ul |
| H2O | 0.4ul |
| MIP pdt (after 100X dilution) | 5ul |
| Total volume | 15ul |

Roche LC480 qPCR protocol

95℃	10min

95℃	10sec

55℃	30sec

72℃	10sec

Goto 2, 45X

40℃	30sec
