## Supplementary material for "Telomere length among Chinese oldest old": Tables TL 14y Os 20220510

**Table 1.** Telomere length (T/S ratio) by 14y characteristics of males and females.

|  | All (n=555) | |  | Males (n=326) | |  | Females (n=229) | |
| --- | --- | --- | --- | --- | --- | --- | --- | --- |
|  | Mean ± SD | P | N | Mean ± SD | P | N | Mean ± SD | P |
| Sex |  | **<0.001** |  |  |  |  |  |  |
| Males | 0.97±0.20 |  |  | -- | -- |  | -- | -- |
| Females | 1.07±0.18 |  |  | -- | -- |  | -- | -- |
| Age, years |  | 0.639 |  |  | 0.968 |  |  | 0.947 |
| 75-79 | 1.00±0.19 |  | 52 | 0.97±0.21 |  | 25 | 1.05±0.15 |  |
| 80-84 | 1.01±0.20 |  | 167 | 0.97±0.20 |  | 104 | 1.09±0.18 |  |
| 85+ | 1.01±0.20 |  | 107 | 0.97±0.22 |  | 100 | 1.05±0.17 |  |
| Education level |  | **0.016** |  |  | 0.247 |  |  | 0.681 |
| Primary or below | 1.03±0.19 |  | 169 | 0.98±0.20 |  | 183 | 1.07±0.18 |  |
| Secondary or above | 0.98±0.21 |  | 157 | 0.96±0.21 |  | 46 | 1.08±0.19 |  |
| SES ladder – Community |  | 0.156 |  |  | 0.104 |  |  | 0.615 |
| 1-3 | 1.02±0.19 |  | 63 | 0.97±0.18 |  | 42 | 1.08±0.17 |  |
| 4-5 | 1.02±0.20 |  | 171 | 0.99±0.21 |  | 125 | 1.06±0.19 |  |
| 6-10 | 0.98±0.20 |  | 83 | 0.92±0.20 |  | 58 | 1.07±0.16 |  |
| SES ladder – Hong Kong |  | 0.738 |  |  | 0.795 |  |  | 0.486 |
| 1-3 | 1.01±0.20 |  | 141 | 0.97±0.20 |  | 110 | 1.06±0.18 |  |
| 4-5 | 1.01±0.21 |  | 128 | 0.97±0.21 |  | 93 | 1.07±0.18 |  |
| 6-10 | 1.00±0.20 |  | 54 | 0.97±0.21 |  | 24 | 1.09±0.14 |  |
| Smoking status |  | **0.007** |  |  | 0.086 |  |  | 0.072 |
| Never/ex-smoker | 1.01±0.20 |  | 315 | 0.97±0.21 |  | 227 | 1.07±0.18 |  |
| Current smoker | 0.86±0.14 |  | 11 | 0.87±0.15 |  | 2 | 0.85±0.13 |  |
| Current drinker |  | 0.241 |  |  | 0.710 |  |  | 0.198 |
| No | 1.01±0.20 |  | 281 | 0.97±0.20 |  | 224 | 1.07±0.18 |  |
| Yes | 0.98±0.20 |  | 45 | 0.98±0.21 |  | 5 | 0.97±0.11 |  |
| Physical activity level |  | 0.084 |  |  | 0.256 |  |  | 0.282 |
| Q1 | 1.04±0.21 |  | 60 | 0.99±0.22 |  | 72 | 1.08±0.20 |  |
| Q2 | 0.99±0.20 |  | 78 | 0.96±0.20 |  | 58 | 1.03±0.19 |  |
| Q3 | 1.04±0.20 |  | 77 | 1.02±0.22 |  | 57 | 1.07±0.15 |  |
| Q4 | 0.98±0.19 |  | 101 | 0.93±0.18 |  | 36 | 1.11±0.17 |  |
| Low BMI - No | 1.02±0.20 | 0.152 | 282 | 0.98±0.21 | 0.058 | 197 | 1.07±0.18 | 0.958 |
| Yes | 0.98±0.19 |  | 42 | 0.91±0.17 |  | 30 | 1.07±0.17 |  |
| No. of chronic diseases |  | 0.163 |  |  | 0.250 |  |  | 0.266 |
| 0-1 | 1.00±0.19 |  | 178 | 0.96±0.20 |  | 132 | 1.06±0.17 |  |
| ≥2 | 1.03±0.21 |  | 137 | 0.99±0.22 |  | 91 | 1.08±0.18 |  |
| Diabetes – No | 1.01±0.20 | 0.453 | 265 | 0.96±0.21 | 0.397 | 182 | 1.07±0.17 | 0.950 |
| Yes | 1.02±0.20 |  | 61 | 0.99±0.20 |  | 47 | 1.07±0.19 |  |
| Stroke – No | 1.01±0.20 | 0.333 | 268 | 0.96±0.20 | 0.272 | 206 | 1.06±0.18 | 0.177 |
| Yes | 1.03±0.21 |  | 58 | 1.00±0.23 |  | 23 | 1.12±0.14 |  |
| CVD – No | 1.01±0.20 | 0.682 | 231 | 0.97±0.20 | 0.653 | 181 | 1.06±0.18 | 0.326 |
| Yes | 1.00±0.21 |  | 94 | 0.96±0.22 |  | 46 | 1.09±0.17 |  |
| Cognitive impairment |  | 0.303 |  |  | 0.789 |  |  | 0.361 |
| No (MMSE≥24) | 1.01±0.20 |  | 276 | 0.97±0.20 |  | 144 | 1.08±0.18 |  |
| Yes | 1.03±0.19 |  | 50 | 0.98±0.21 |  | 85 | 1.05±0.18 |  |
| Frailty status |  | 0.218 |  |  | 0.315 |  |  | 0.094 |
| Robust | 0.97±0.20 |  | 33 | 0.93±0.19 |  | 8 | 1.17±0.14 |  |
| Pre-frail | 1.01±0.20 |  | 200 | 0.98±0.20 |  | 104 | 1.08±0.16 |  |
| Frail | 1.01±0.20 |  | 93 | 0.97±0.21 |  | 117 | 1.05±0.19 |  |
| Sarcopenia – No | 1.01±0.19 | 0.525 | 137 | 0.95±0.18 | 0.114 | 137 | 1.06±0.18 | 0.637 |
| Yes | 1.02±0.21 |  | 164 | 0.99±0.22 |  | 86 | 1.07±0.18 |  |

SD, standard deviation; SES, subjective socioeconomic status; BMI, body mass index; CVD, cardiovascular diseases; MMSE, Mini-Mental State Examination

**Table 2.** Crude and adjusted associations between telomere length (T/S ratio) and prevalent frailty and sarcopenia status^a^

|  | Unadjusted OR | 95% CI | P | Adjusted OR | 95% CI | P |
| --- | --- | --- | --- | --- | --- | --- |
| *Frailty* |  |  |  |  |  |  |
| Males | 0.99 | 0.31-3.23 | 0.990 | 0.55 | 0.10-3.06 | 0.490 |
| Females | 0.31 | 0.07-1.37 | 0.122 | 0.19 | 0.02-1.51 | 0.117 |
| *Sarcopenia* |  |  |  |  |  |  |
| Males | 2.45 | 0.79-7.59 | 0.120 | 4.07 | 0.96-17.3 | 0.057 |
| Females | 1.45 | 0.31-6.74 | 0.635 | 0.74 | 0.10-5.62 | 0.769 |

OR, odds ratio; CI, confidence intervals.

Adjusted for age, education level (primary or below/secondary and above), body mass index, socioeconomic status Hong Kong ladder and community status ladder, number of chronic diseases, current smoker (yes/no), current drinker (yes/no), PASE score and cognitive impairment (yes/no).

^a^Fried’s criteria: no energy, HGS ≤first quintile, 6-m walking speed ≤first quintile, physical activity level ≤first quintile, weight loss ≥5% in past 6 months.
